# Facilitators and barriers to using iris scanning for identification of mobile populations at high risk of HIV in fishing communities of Lake Victoria in East Africa

**DOI:** 10.1101/2025.09.30.25337025

**Authors:** Esther Awino, Sarah Nakamanya, Ubaldo Bahemuka, Joseph Ssembatya, Freddie Mukasa Kibengo, Zacchary Kwena, Elialilia Okello, Gertrude Nanyonjo, Brenda Okech, Ali Ssetaala, William Kidega, Eugene Ruzagira, Elizabeth Bukusi, Saidi Kapiga, Matt A. Price, Patricia Fast, Janet Seeley

## Abstract

The growing concern over participant identification in research, spurred on by flaws in conventional methods like name/date of birth-based identity cards, hospital cards and names, has led to adverse consequences, such as drug misadministration, co-enrolment and missed appointment visits. This study examines the facilitators and barriers associated with implementing biometric iris scanning for participant identification among mobile women and men working in the fishing communities of Lake Victoria. We aim to address challenges in participant identification and contribute to improved research practices.

Twenty-four focus group discussions (FGDs) were conducted between 2021 and 2022 across Uganda, Kenya and Tanzania, involving participants aged 18 years and above. The study specifically targeted members of mobile fishing communities residing along the shores of Lake Victoria. The participating research organizations, all partners of the Lake Victoria Consortium for Health Research (LVCHR), operated at the four sites: Masaka and Entebbe in Uganda and Mwanza in Tanzania and Kisumu in Kenya. Each site covering two fishing communities. Participants had an experiential session with the iris camera/scanner, voluntarily undergoing the scanning process before participating in a subsequent FGD. Data collected from the FGDs were transcribed, coded, and subjected to inductive thematic analysis to identify themes directly emerging from the participants’ narratives. This methodology was employed to gain insights into the participants’ perceptions and experiences of the implementation of iris scanning technology in these communities.

Overall, there was a broad acceptance of the biometric iris scanning technology, attributable to the comprehensive sensitization efforts among those who participated in the research, effective communication, and the provision of clear information regarding the intentions behind its introduction. However, because the participants had never been exposed to this technology before, various misconceptions, expectations, fears and concerns regarding privacy and safety emerged. These could potentially act as barriers to its adoption in the wider community looking beyond research participants.

There is need to provide comprehensive sensitization and health education through communication channels tailored to the needs of specific communities, while engaging them is key in guiding sensitization procedures and improving acceptability. These approaches will help increase acceptability and reduce hesitancy and fears associated with the use of new technologies, such as the iris scanner.

## Introduction

Due to the limitations of conventional participant identification methods, researchers in low- and middle-income countries are gradually transitioning to advanced technologies for accurate identification of individuals (1). This shift is particularly critical for mobile populations, such as fishing communities, where inaccurate participant identification can pose significant challenges (2). Men and women in such communities, engaged in related employment who move for work, are a priority for researchers for devising solutions to ensure accurate and reliable participant recognition (3).

Traditional identification methods, including identity cards, hospital cards, names, dates of birth, and fingerprints, have proved to be inadequate and fraught with challenges. Issues related to most of the above mentioned traditional methods such as, mismatch of patients to their drugs, impersonation, loss of identity cards or hospital cards have hampered the execution of research activities, leading to errors in health research, such as administering incorrect drug doses as a result of mismatching patients to drugs (4). Utilization of biometrics, particularly iris scanning, presents a promising solution to mitigate or eliminate the irregularities associated with traditional identification methods (5).

A biometric is a measure of identity based on physiological (finger print, face, eye iris or retina) or behavioral (speech, signature) characteristic (6). The iris scanner in particular employs unique personal identifiers derived from the eye, assigning a personalized identifier to each individual while maintaining confidentiality about other personal characteristics. This ensures quick and error-free information retrieval (3, 7). Demonstrating high reliability and accuracy, iris scanning has gained widespread acceptance in various sectors, including healthcare, security, and industries (8–10). The use of biometric iris scanning for identification is rapidly growing in resource-constrained settings including in sub-Saharan Africa. For example, studies on the use of iris scanning for identification in the Democratic republic of Congo and Kenya have shown considerable evidence of acceptability, accuracy and feasibility in these settings (2–4, 8, 10). However, successful use of the technology will require careful consideration of the potential barriers and facilitators to improve uptake and use.

Our study examined the barriers and facilitators to using iris scanning for identifying mobile populations at high risk of acquiring HIV in fishing communities around Lake Victoria in Uganda, Kenya and Tanzania, East Africa.

## Methods

### Study design and setting

This multi-site qualitative study was conducted by partner organizations of the Lake Victoria Consortium for Health Research (LVCHR), a collaborative alliance of independent health research institutions dedicated to investigating health-related issues within the fishing communities of Lake Victoria. LVCHR partner organizations include the Kenya Medical Research Institute (KEMRI) in Kenya, the Mwanza Intervention Trials Unit (MITU) in Tanzania, the Uganda Virus Research Institute-International AIDS Vaccine Initiative (UVRI-IAVI) in Uganda, and the Medical Research Council/Uganda Virus Research Institute and London School of Hygiene and Tropical Medicine (MRC/UVRI & LSHTM) Uganda Research Unit also based in Uganda. The participating research organizations operated at the four sites: Masaka and Entebbe in Uganda and Mwanza in Tanzania and Kisumu in Kenya. Each site covered two fishing communities.

## Sampling and data collection

Data were collected between March and June 2021. A total of 24 focus group discussions (FGDs) were conducted, 8 from each of the 3 countries.

Participants were mobilized through their community leaders (VHTs, chairpersons) and given information about the planned study in their community, including the fact that the research involved the use of an iris scan for identification. The leaders aided in identifying participants who were eligible to participate in the study.

Participants were selected purposively to include different groups of people. The groups were stratified by age and gender to ensure perspectives from both men and women across the different age groups. Each site conducted four group discussions with younger participants (18-24 years, male and female) and four group discussions for older participants (35 years and above, male and female). Each group comprised of 6 to 8 participants.

Prior to each group discussion, the functionality of the technology was explained and demonstrated during the informed consent process. Each participant was given an opportunity to look through the iris scanner, which the teams always carried along for the demonstration purpose. The participants were told that the iris scan captured only the image of the iris, then it generated a unique serial number for each of them. This iris scan was connected to a computer that stored the data to a folder on the server of each organization in accordance with the data protection policy of these organizations. Only those involved in this research had access to these data.

During group discussions, participants were asked whether they had seen or heard of an iris scanner before and to share their opinions on iris scanning as an identification method. They were also asked their main concerns about its use, effective ways for engaging and encouraging participation in a planned study to assess the feasibility its use for identification of research participants, and potential barriers that could hinder study participation.

Each focus group discussion was facilitated by two experienced social science research assistants, with one serving as the moderator and the other as the note taker. All group discussions took place at the fishing sites in a quiet and convenient location chosen by study participants.

Group discussions were conducted in the local languages - Luganda in Uganda, Kiswahili in Tanzania and Dholuo in Kenya. Discussions followed a semi-structured topic guide and were audio recorded, with each session lasting an average of 45 minutes.

### Data management and analysis

Audio recordings from FGDs at each site were transcribed and translated from the local languages into English. FGD recordings and transcripts were labelled using identification numbers and saved on password protected computers with access only allowed to study teams. To enhance confidentiality, data were anonymized and each transcript assigned a unique identification number.

The typed transcripts were reviewed by the lead researchers at each site to ensure data completeness. Any inconsistencies or missing information were sent back to the research assistants for clarification.

The analysis involved a preliminary review by the same researchers who collected the data at each site. Working with the lead researcher, they identified common codes, as well as initial themes and patterns emerging from the data. These codes were shared with the other teams from the different sites, discussed and agreed upon. A coding matrix was then developed and shared with other team members for refining. This was then finalized to one codebook that was used at all sites to complete the coding exercise. Similar codes were grouped together and these broader categories or themes provided insights into barriers and facilitators of using iris scanning for identification.

### Ethical approval

Ethical approval was obtained in each country from the appropriate regulatory body: the Uganda Virus Research Institute Research Ethics Committee (UVRI REC # 605) and Uganda National Council for Science and Technology (UNCST # 4470) for Uganda; KEMRI Scientific and Ethics Review Unit (SERU # 3593) for Kenya; the National Health Research Ethical Committee (NatHREC # MR/53/100/637 & 659) for Tanzania and the London School of Hygiene and Tropical Medicine Ethics Committee (LSHTM # 22449 & 22639). All participants provided written informed consent before taking part in the study and were compensated for their time at the end of their participation.

### Results

A total of 156 individuals participated in FGDs across the sites: MRC/UVRI and LSHTM and UVRI/IAVI (54), KEMRI (50) and MITU (52). Of these, 35 were older men (≥35 years) and 36 older women (≥35 years while 36 were young men (18-24 years) and 48 were young women.

Overall, most of the participants had attended primary school or a few years of secondary school education. Only a small number had completed secondary school. The majority were engaged in fish-related work, while others operated non fish- related small businesses which served the fishing community. Some participants, particularly women, reported having no paid employment and were primarily involved in unpaid domestic work.

**Table 1.** Participant’s demographics. *Missing some data on marital status, but available data shows majority were single/separated

| **SITES** | **MRC/UVRI&LSHTM**<br>**& UVRI-IAVI**<br>**(Uganda)** | **KEMRI**<br>**(Kenya)** | **MITU**<br>**(Tanzania)** | **Total** | **%** |
| --- | --- | --- | --- | --- | --- |
| **Variables** |  |  |  |  |  |
|  | #54 | #50 | #52 | #156 |  |
| **Sex** |  |  |  |  |  |
| Male | 26 | 20 | 26 | 72 | 46% |
| Female | 28 | 30 | 26 | 84 | 54% |
| **AGE** |  |  |  |  |  |
| 18-24(females) | 16 | 20 | 12 | 48 | 31% |
| 35+(females) | 12 | 10 | 14 | 36 | 23% |
| 18-24(males) | 14 | 10 | 13 | 36 | 23% |
| 35+(males) | 12 | 10 | 13 | 35 | 22% |
| **Education** |  |  |  |  |  |
| Primary | 31 | 33 | 37 | 101 | 65% |
| secondary | 23 | 17 | 15 | 55 | 35% |
| **Occupation** |  |  |  |  |  |
| Fish related work | 31 | 20 | 38 | 89 | 57% |
| No fish related<br>businesses | 10 | 10 | 03 | 23 | 15% |
| other | 13 | 20 | 11 | 44 | 28% |
| **Marital status** |  |  |  |  |  |
| Married/cohabiting | 18 | 17 | * |  |  |
| Separated/single | 36 | 33 | * |  |  |

### Facilitators

These factors, as highlighted by participants, would potentially contribute to the acceptability of the iris scanner and facilitate its use in the target communities.

### Awareness and education

Many of the participants suggested that for the iris scanner to be readily accepted as a participant identification tool in health research, comprehensive sensitization and clear communication about the technology would be essential. This should be done through media channels such as radio, television, social media, as well as through persons who communities typically trust, listen to and confide in such as local leaders, church leaders, teachers, and health workers. Participants noted that effective sensitization would enhance the communities’ understanding of the new technology and its purpose, which would in turn increase its uptake and use.

> “…something to do with mass education, it helps so much in the community; you can involve the chiefs, radio discussing about how it works and its benefits, at least they can have a clue about it, so when you approach one, they will have heard it over the radio. (young men, Kenya)

> *“Others will be afraid, but this can be resolved with educating the community so that they understand the importance of iris scanner, and that there is no any harm they will get for using the device” (older women, Tanzania)*.

Other participants suggested that highlighting the positive aspects including expected benefits of the technology would encourage its uptake, not solely for research purposes. For instance, if people get to know that it would help improve service delivery in the health facilities other than just research by reducing delays and bureaucracies, they would be more willing to welcome its use. In other words, participants had expectations for this iris scan and in many instances, they would explain themselves in reference to “if” it was adopted in health facilities where they access healthcare. One participant shared this perspective, stating:

> *“I may not individually have any fears about using an iris scanner because I have looked at it in a different perspective, in regard to improving health care services delivery but some other people may look at it differently” (Older women, Uganda*).

> *“From your information, the iris scanner is a good device because it makes it easier to identify. I might have forgotten my ID at home, and I cannot find it at the time, so this eye scanner helps to identify a person when they need service without a need to register again.” (young men, Tanzania)*

Participants also suggested training community health educators on the use of the technology who could in turn educate and inform their communities. They viewed this as another strategy that could facilitate the uptake of this new technology.

> *“I advise that the experts of that device call a meeting with street leaders and educate them and these in turn educate their people or the experts can also use young people from the respective streets to help educate people because the local communities would trust people that they know.* (*Young men, Tanzania)*

Overall, sensitizing communities on the use and benefits of iris scanning was mentioned as being key to promoting its acceptability and use.

### Ease, convenience and timing

Participants viewed the technology as a convenient solution to the challenges of the burden of carrying and presenting traditional forms for identification. They noted that study participants sometimes missed appointments simply because they had lost their appointment cards or left them elsewhere. In addition, they highlighted that the iris scanner would save time by eliminating the need for a researcher’s manual search through files to identify participants or retrieve their details.

> *“…it would have been a great idea to use an iris scanner because it would reduce on the bureaucratic tendencies/ delays in health centers since in the first capture of the iris, all patient details are recorded, the next time they go back, there would be no delays.” (older women, Uganda)*.

> *“It makes work easy when we come back, they can identify those who had participated before.”(young women, Kenya)*

Participants also highlighted the advantages of iris scanning over other methods like cards and fingerprint identification, noting that fingerprints can become difficult to detect due to worn out ridges caused by certain types of work.

> *“I support the use of iris scanner because sometimes the use of traditional way such as finger print has failed because for people who work in fishing industries their finger scratches therefore it becomes hard for the finger scanner to detect” (young men, Tanzania)*.

They pointed out, however, that although they perceived the technology to be convenient, its successful use was dependent on the availability of study participants. For example, researchers needed to clearly understand the seasonal fishing patterns, which impact their work routines and availability. To maximise the benefits from the technology, researchers should align study activities with participants’ availability, ensuring engagement at the most suitable times for these mobile communities.

> *“Time factor is important, in this community there is a season where people are very busy that you hardly see anyone and there are other times especially during the high moon which lasts for about 7 days, where they are relaxing from work and it could be used to explain the new technology that we intend to bring.” (young men, Uganda)*

### Involving communities and other key stakeholders

Participants emphasized that involving community leaders would be crucial in encouraging individuals to participate in research studies evaluating new technologies. These leaders play a big role in fostering trust and building confidence within the community.

> *“It is important to engage schools and churches to help to inform people about the use of iris scanning. Churches and schools are the place where many people come together therefore it will help in sensitizing and mobilizing people.*” (*young women, Tanzania)*

> *“Also, involving the community leaders need to be considered in these study initiations so as to build confidence and trust in people that they can be part of the study” (older women, Uganda)*.

Some participants felt that because iris scanning was a new concept in the community, those who experienced it firsthand in the FGDs should share their experiences and the benefits of using the technology with others. They believed that these personal testimonies would help encourage others to adopt the technology and participate in future studies.

> *“I would suggest that we can convince others if they happen to see how we might have benefitted from the study, and if they publicly show what the benefits are from this study, in that case many more youth will be participating” (young men, Uganda)*.

> *“We’ve participated, been trained and informed about the benefits, so as to go tell those who haven’t participated, we can teach them on the benefits, encouraging them that’s how they’ll get the courage to come.” (young women, Kenya)*.

In summary, research participants thought iris scanning was useful, reliable, and efficient compared to traditional identification techniques. It saves time, eliminates the need for paper ID cards, and reduces bureaucratic hold-ups. Additionally, iris scanning was thought to be a more reliable method than fingerprint identification, which can be affected by physical and environmental changes. However, participants noted that it was important for researchers to engage the communities as well as consider the most appropriate timing for their targeted populations so as to easily implement and benefit from the use of this technology.

### Barriers

In this context, barriers refer to factors that may contribute to an individual’s refusal to take part in a study involving the use of iris scan in their communities, as well as challenges associated with usability of the technology.

### Safety and privacy concerns

As iris scanning was a new technology for many, most participants expressed fears and concerns, particularly regarding its potential impact on physical health. The fears and concerns derived mainly from the limited understanding of the technology. Across all sites, participants feared that testing this new technology could harm eyesight, especially among older people whose sight was already deteriorating. Additionally, the camera’s flashlight and the repeated use of the device from one person to another raised concerns about potential eye damage and the risk of transmitting eye infections.

> *“I suspect the camera does not only take the iris but also could penetrate the brain and damage it. I suggest that there is need to give better explanation about the iris scanner. The camera may make me go blind as a side effect.” (Older women, Uganda)*

> *“The thing that can make a person not participate in the research is fear, a person may think that the device is a source of diseases or a way to transplant diseases or can cause blindness.” (young men, Tanzania)*

Participants expressed their misconceptions about the iris scan for instance, that it was a disguised way of making the iris scan mandatory like it was for the case of national identity cards before for the case of Uganda. Thus, their participation to cooperate was driven by the perception that using the iris scan may be a compulsory measure in future. Others thought it was a disguise for COVID-19 vaccination for those who had refused to get vaccination at the time of the study. Below are some of the participants’ narratives:

> *“…it’s better for people to cooperate and understand what is being brought to them, there is a time they brought registration of national IDs and people were reluctant. This brought many issues with time as National IDs became mandatory, so maybe same case for the iris scan.” (young women)*

> *“…some may hesitate to participate because of fearing that it may be a way of vaccinating COVID to people who do not want vaccination.” (FGD young women, Tanzania)*

Given these anticipated side effects, participants questioned the safety of iris scanning. They also feared that the iris scanner could impair their vision, potentially leading to blindness.

Additionally, they worried that, if adverse effects occurred, there would be no guarantee of receiving treatment. Moreover, they believed that these concerns would be significant within the broader community since they have little trust in their governments.

> *“… they will refuse to participate in the study because they have that inner doubt in researchers that some of them don’t tell the truth about their studies… an example is the Covid-19 vaccine, some of the vaccines can damage the heart and liver and that is why we cannot just accept anything brought by the government, it’s a matter of being cautious.” (young women, Uganda)*.

Participants also expressed concerns about the potential misuse of their personal information, particularly the possibility of it being shared with authorities such as police or other law enforcement institutions through research organizations or hospitals. Most of the participants expressed the fear that individuals with a history of legal issues might be identified and targeted if their credentials were recorded.

Some participants expressed the fear that the introduction of the iris scanner could have hidden political motives that might negatively impact them. They viewed this as a serious barrier to acceptance of the technology, particularly if the communities were not well sensitized and given clear information on its purpose:

> *“Insufficient information about iris scanning and its benefits can also cause people to hesitate in participating in the process, the belief that people may have about the possibility of the state hiding behind research to track down criminals through the use of iris scanners can lead them not to participate especially people with criminal background who came in the fishing communities to hide.”(young women, Tanzania)*

> *“If it was during campaign, we would automatically think xx (name withheld) has brought the mechanism of looking through the camera to make ensure he is elected. The only problem in* that *is that some people think that you are conmen who have come with hidden intentions.”*

(*young women, Uganda)*

Despite clear explanations and demonstrations provided by the researchers on how the scanner functions regarding research activities, some participants had concerns about the motives behind the introduction of the iris scanner to their communities. They mentioned that any issues experienced by individuals after using the scanner could be attributed to its use.

> *“The only problem is that some people think that you are conmen who have come with hidden intentions. There are very many conmen who lie to people, there are people who come to the community with hidden intentions including health workers and this may stop people from being part of a study” (young women, Uganda)*.

As a result of what they termed as being too little information provided by the team and their perceptions of a ‘hidden agenda’, participants expressed concerns about how the information gathered was going to be used. In these group discussions, some of them constantly associated iris scanning to law enforcement agencies. There was fear that the scanned data were probably going to be used or leaked to the government or law enforcement agencies to track criminals instead of health research activities.

> *“They can be afraid, like the machine scans the iris, one can ask themselves where you want to take the scanned iris, this can prevent one from participating in the study.” (Young women, Kenya)*

> *“Some people will not participate fearing that one day they can commit a crime and therefore, it will be easier for them to be traced because information captured through the iris scanner is unique and it can be shared easily across the country.” (Young women, Tanzania*)

Additionally, participants, especially at the Ugandan site, expressed concerns that the timing of the iris scanner’s introduction amidst ongoing political unrest was poor. They feared that community members might perceive it as a tool for identifying political opponents for arrest.

> *“The country is in a political turmoil besides COVID-19, so people in the community may think that the Iris scanning is political and that the government may have a hidden agenda, the differences in politics* (political differences) *may affect people’s participation.” (Older women, Uganda)*

Participants noted that not everyone in their communities was educated, and some, particularly the elderly, might be skeptical of the iris scanner, even fearing that the researchers intend to harm them and are not always transparent. There was a concern that certain individuals might associate the technology with negative or supernatural connotations, such as links to a “Chip” that could be used to track their movements. However, they believed that with proper education, these concerns could be addressed, and the community would ultimately accept the technology.

> *“That is what some people will actually think, they will refuse to participate in the study because they have that inner doubt in researchers that some of them don’t tell the truth about their studies.” (Young women, Uganda)*

> *“ […] we have those who don’t welcome the idea of using machines, they may think that maybe you are capturing their eyes and go use it somewhere else” (young women, Kenya)*.

So, these fears expressed in participants’ narratives, which included the fear of side effects and spread of infectious diseases together with the suspicion of a hidden agenda and a fear for their privacy and confidentiality were among the barriers to the use of iris scanning.

### Limited or no personal benefits /gains

While the introduction of the iris scanner in the conduct of research was welcomed and largely met with enthusiasm, a big number of participants expressed challenges in understanding the tangible ways in which they would benefit as individuals from its implementation. Much as the technology was seen as innovative and useful, many participants lacked a clear understanding on its immediate relevance to their own lives. For example, some individuals thought that the iris scanner would be able to detect or diagnose and treat eye-related health complications. Others simply wanted to know how their participation would result in more tangible benefits, particularly monetary benefits.

> *“Me I don’t see the benefit of the iris scanner to us rather we have not understood the significance of the iris scanner on their lives.” (Young women, Uganda)*

> *“…it will be a good idea to use the iris scanner if it was going to also detect a medical condition…” (Young women, Uganda)*

So, an important concern from participants’ narratives was the economic reality that participants and many other people in their communities faced. They indicated that people are often preoccupied with their daily activities, particularly the work that provides their livelihood. In this respect, taking time away from income-generating activities to engage in a research study, especially one whose benefits are not obvious, was seen as a challenge that could deter many from participating. Therefore, without any form of compensation or incentive, participants felt it would be unrealistic to expect individuals to forego their personal or economic obligations and participate in a research study.

> *“We are different people and everyone is busy, there is no one who will spend their time on such useless things that do not benefit them, people are busy in different things that earn them income, most people won’t come and spend their time here and they leave with nothing at all.” (Young men, Kenya)*

> *“Many people are currently involved in business, so it will be difficult for people to close their business and participate in this activity; maybe if you will compensate them with even two thousand shillings each will motivate each other.” (Older women, Tanzania)*.

Overall, participants were skeptical about the direct benefits of the iris scanner to their lives and mentioned that these expectations influence their decisions to take part in such studies. Therefore, lack of perceived benefits (direct or indirect) may deter some from participation.

## Discussion and recommendations

This study examined the perceived barriers and facilitators that influenced use of using iris scanning for identification to prevent double enrolment of mobile research participants in fishing communities around Lake Victoria. Our findings revealed a complex interplay of factors that influenced use of the technology, acting either as enablers/facilitators or hindrances/barriers to its use. A key factor perceived to facilitate use of the technology was awareness creation. The novelty of iris scanning in the community increased participants’ skepticism, as they lacked prior information about the technology.

Moreover, similar studies have shown that a lack of information or awareness can hinder acceptability in many communities (2, 4) and, as suggested by our participants, educating communities has been identified in other studies as critical factor in raising awareness and ensuring the success of health research (11, 12).

Using the different available communication channels that are most trusted by the community is crucial when introducing new technologies or products. To reduce mistrust and foster confidence in the iris scanning technology, it is essential to educate communities through transparent communication and active engagement. Existing literature demonstrates that involvement of community stakeholders fosters a sense of ownership, accountability, increased awareness, transparency, and informs the design of interventions, ultimately increases acceptability while alleviating fears and concerns among end users (13, 14).

Similarly, participants in our study emphasized the importance of involving communities and trusted community figures to build trust and confidence in iris scanning technology.

This approach could positively influence the community’s perception of iris scanning(1). It not only helps to reduce hesitancy but also addresses misconceptions, misinformation, doubts, and fears, as highlighted in participants’ narratives regarding what could facilitate or enhance the uptake of iris scanning for identifying research participants (1, 2). Other studies have shown that community leaders play a significant role in implementing policies and influencing their communities, as people tend to trust and look up to them for guidance (1, 13). So, community involvement could help address potential resistance while improving acceptance of the iris scanning technology by ensuring that its implementation aligns with community expectations.

Despite the facilitators, barriers to adoption of iris scanning were identified and these were mainly about privacy concerns. Fears were expressed about data misuse, surveillance, and unauthorized access to their biometric information. Studies conducted in fishing communities around Lake Victoria have shown how these areas serve as hideouts not only for criminals but also individuals seeking to forge new identities (15). In such contexts, concerns about privacy and safety could hinder participants’ willingness to adopt iris scanning technology as they had it at the back of their minds that accepting this technology in their setting would compromise their personal information. Thus they worried that the data might be linked to law enforcement agencies rather than being used solely for health research purposes. (16). Other privacy safeguards and measures like data protection policies and informed consent processes should be conducted in the best way possible to ensure maximum comprehension of the processes, including privacy (16–18). In one study that explored health worker perceptions and user experiences in biometric technology in Zambia, it was recommended that high level trainings be undertaken to ensure data integrity as a way of avoiding breach and maximizing benefits (18).

The finding that some participants did not perceive the iris scanning technology as beneficial is concerning, as it could significantly hinder its adoption. Participants’ questions about how the technology would directly benefit them, especially when the research outcomes were not expected to yield tangible improvements in their health or well-being poses an obstacle to its use. The lack of clear individual benefits was foreseen as a deterrent to the technology adoption. Clear and effective communication, including raising awareness of the indirect benefits of iris scanning, is essential. The misconception that iris scanning detects or treats eye defects, also reported in other studies (19, 20), should be actively addressed to improve understanding and acceptance.

From the discussions, it was evident that people are less inclined to participate in research activities if they cannot foresee clear, direct, and personal benefits. This highlights the importance of effectively communicating the purpose and potential advantages of new technologies in research, as well as considering practical strategies such as incentives or reimbursements to encourage wider community engagement.

Therefore, the effective adoption of iris scanning technology for identifying mobile research participants in fishing communities around Lake Victoria requires comprehensive awareness efforts and proactive community engagement. Engaging with communities through transparent communication and health education, involving trusted community leaders, and emphasizing the technology’s advantages are crucial. Similarly, addressing concerns about safety, privacy, and personal benefits requires robust data governance policies as well as offering transparent benefits.

## Limitations of the study

One of the limitations of our study was that the use of the iris scanner was limited to a one-off demonstration before the discussions, which did not allow participants adequate time to experience the technology before giving their opinions. Participants’ experiences were based on sample demonstrations designed to give participants a basic understanding of the scanner before participating in the study. Consequently, participants’ views may differ from those formed through more extensive experience with the technology.

Secondly, in addition to the small sample, the study was conducted in fishing communities in remote areas, where exposure to and experience with the technology may have been limited. Consequently, our findings may not be generalizable to other communities within the region. However, the high mobility of this population made it a suitable group for testing the technology and in the use of focus group discussions to collect data. These allow for in-depth exploration of attitudes, beliefs, and social norms related to the use of iris scanning technology. They encourage participants to share and build on each other’s experiences, revealing nuanced barriers and facilitators of using the technology.

## Conclusion

Several factors were perceived to impede the use of iris scanning for identification of highly mobile research participants from communities around Lake Victoria. Many of these challenges stemmed from the novelty of the technology and the limited information regarding the new technology. To ensure the successful adoption of iris scanning in these communities, a balanced approach that acknowledges both barriers and facilitators is needed. Key facilitators may include ongoing awareness campaigns, educational programs, continuous user support, and community involvement in the development and testing of the technology.

## Data Availability

The qualitative data underlying and illustrating the findings of the study we collected and synthesized are presented within the manuscript. Anonymized transcripts are not publicly available due to ethical and legal reasons as they contain information that could potentially compromise participant privacy. Data requests may be sent to.

## Acknowledgements

We acknowledge the Leadership the Lake Victoria Consortium for Health Research (LVCHR) and the collaboration and support of its partner members. Special thanks to; Jan De Bont and the entire IAVI leadership who supported funding of this work. We thank the study field team members at the different sites; Edmund Kisanga, Derick Deogratias, John Luwayi, Bernard Dajo, Catherine Makokha, George Ochieng, Seth Owino, Geofrey Basalirwa, Mathias Wambuzi, Henry Kaloma, Patricia Mulungi, Marion Namuleme, Tom Wambi and Joel Odenyo for their invaluable effort in implementing the field work. More importantly, we thank all study participants for agreeing to provide information for the study and the community gatekeepers for their support.

## Author contributions

Conception and design of the study: PF, ZK, JS, EO, SK, BO, FMK, SK, EB, AS, UB, WK, ER, MP; Study implementation: EO, ZK, BO, GN, SN, JS, AS, FMK, UB; Writing original draft: AS, SN, UB, GN, EO; Critical revision of the manuscript for important intellectual content; JS, ZK, BO, AS, FMK, WK, EB, PF, SK, ER, MP. All authors read and approved the final manuscript.

## References

1. Cambaco O, Gachuhi N, Distler R, Cuinhane C, Parker E, Mucavele E, et al. Acceptability and perceived facilitators and barriers to the usability of biometric registration among infants and children in Manhiça district, Mozambique: A qualitative study. Plos one. 2021;16(12):e0260631.

2. Okello E, Ayieko P, Kwena Z, Nanyonjo G, Bahemuka U, Price M, et al. Acceptability and applicability of biometric iris scanning for the identification and follow up of highly mobile research participants living in fishing communities along the shores of Lake Victoria in Kenya, Tanzania, and Uganda. International Journal of Medical Informatics. 2023;172:105018.

3. Zola Matuvanga T, Johnson G, Larivière Y, Esanga Longomo E, Matangila J, Maketa V, et al. Use of iris scanning for biometric recognition of healthy adults participating in an Ebola vaccine trial in the Democratic Republic of the Congo: mixed methods study. Journal of Medical Internet Research. 2021;23(8):e28573.

4. Njoroge A, Dunbar MD, Abuna F, Simpson P, Macharia P, Betz B, et al. Feasibility and acceptability of an iris biometric system for unique patient identification in routine HIV services in Kenya. International Journal of Medical Informatics. 2020;133:104006.

5. Mwapasa M, Gooding K, Kumwenda M, Nliwasa M, Kaswaswa K, Sambakunsi R, et al. “Are we getting the biometric bioethics right?”–the use of biometrics within the healthcare system in Malawi. Global Bioethics. 2020;31(1):67–80.

6. Giné X, Goldberg J, Sankaranarayanan S, Sheerin P, Yang D. Use of biometric technology in developing countries. In: Cull RJ, Demirguc-Kunt A, Morduch J, editors. Banking the World: empirical foundations of financial inclusion. Boston, USA: MIT Press; 2012. p. 429-44.

7. Kaur N, Juneja M. A review on iris recognition. 2014 Recent Advances in Engineering and Computational Sciences (RAECS). 2014:1-5.

8. Latman NS, Herb E. A field study of the accuracy and reliability of a biometric iris recognition system. Science & Justice. 2013;53(2):98–102.

9. Patel CD, Trivedi S, Patel S. Biometrics in IRIS technology: A survey. International Journal of Scientific and Research Publications. 2012;2(1):1–5.

10. Forchuk C, Donelle L, Capretz M, Bukair F, Kok J, editors. Evaluating Iris Scanning Technology to Link Data Related to Homelessness and Other Disadvantaged Populations with Mental Illness and Addiction. Smart Homes and Health Telematics, Designing a Better Future: Urban Assisted Living: 16th International Conference, ICOST 2018, Singapore, Singapore, July 10-12, 2018, Proceedings 16; 2018: Springer.

11. Durrance-Bagale A, Marzouk M, Tung LS, Agarwal S, Aribou ZM, Ibrahim NBM, et al. Community engagement in health systems interventions and research in conflict-affected countries: a scoping review of approaches. Global Health Action. 2022;15(1):2074131.

12. Corby PM, Schleyer T, Spallek H, Hart TC, Weyant RJ, Corby AL, et al. Using biometrics for participant identification in a research study: a case report. Journal of the American Medical Informatics Association. 2006;13(2):233–5.

13. Waddington H, Sonnenfeld A, Finetti J, Gaarder M, John D, Stevenson J. Citizen engagement in public services in low-and middle-income countries: A mixed-methods systematic review of participation, inclusion, transparency and accountability (PITA) initiatives. Campbell Systematic Reviews. 2019;15(1-2):e1025.

14. Bovaird T. Beyond engagement and participation: User and community coproduction of public services. Public administration review. 2007;67(5):846–60.

15. Nakamanya S, Okello ES, Kwena ZA, Nanyonjo G, Bahemuka UM, Kibengo FM, et al. Social networks, mobility, and HIV risk among women in the fishing communities of Lake Victoria. BMC Women’s Health. 2022;22(1):555.

16. Kavanagh MM, Baral SD, Milanga M, Sugarman J. Biometrics and public health surveillance in criminalised and key populations: policy, ethics, and human rights considerations. The Lancet HIV. 2019;6(1):e51–e9.

17. Myers J, Frieden TR, Bherwani KM, Henning KJ. Ethics in public health research: privacy and public health at risk: public health confidentiality in the digital age. American Journal of public health. 2008;98(5):793–801.

18. Hamapa AM, Zulu JM, Khondowe O, Hangulu L. Healthcare workers’ perceptions and user experiences of biometric technology in the selected healthcare facilities in Zambia. Discover Public Health. 2024;21(1):47.

19. Zola Matuvanga T. Dealing with challenges in an Ebola vaccine trial in a remote and endemic Ebola setting of the Democratic Republic of the Congo. Antwerp, Belgium: University of Antwerp; 2024.

20. Laux D, Luse A, Mennecke B, Townsend AM. Adoption of Biometric Authentication Systems: Implications for Research and Practice in the Deployment of End-User Security Systems. Journal of Organizational Computing and Electronic Commerce. 2011;21(3):221–45.

